# Correlation study of serum 25 hydroxyvitamin D with peripheral neuropathy in type 2 diabetes

**DOI:** 10.1101/2024.12.18.24319256

**Authors:** Senzhen Chen, Jinfeng Chen, Mengting Huang, Yiling Yan, Shujing Zheng, Nuoqi Chen

**Affiliations:** Zhangzhou Affiliated Hospital of Fujian Medical University; Fujian Medical University

**Keywords:** Serum 25 hydroxyvitamin D, diabetic peripheral neuropathy, type 2 diabetes

## Abstract

**Objective:** To investigate the correlation between serum 25 hydroxyvitamin D (25 (OH) D) and peripheral neuropathy of type 2 diabetes (Diabetic Peripheral N europathy, DPN) and its predictive value for DPN.

**Methods:** The subjects were divided into deficient group, insufficient group and normal group, and the relationship between 25(OH)D and DPN and the correlation of 25(OH)D and DPN with various indicators were analyzed.

**Results:** Comparing the DPN group with the NDPN group, 25(OH)D3(22.10±0.77ng/ml vs 24.45±0.66ng/ml), 25(OH)D total (23.12±0.74ng/ml vs 25.68±0.67ng/ml), BMI, RHR, TG, FT3, BUN between the two groups (P <0.05), 25(OH)D3,25(OH)D total, FT3, BMI, and RHR are risk factors for DPN. The ROC curve showed that the optimal cutoff for 25(OH)D3 predicting DPN in T2DM was 18.85ng/ml (AUC 0.76,95% CI 0.697-8.823) and the optimal cutoff for 25(OH)D predicting T2DM was 19.94ng/ml (AUC 0.765,95% CI 0.703-0.828).

**Conclusion:** 25(OH)D can be used as a simple and effective screening method to predict the occurrence of DPN for total 25(OH)D<19.94ng/ml or 25(OH)D3<18.85ng/ml.

## 1. Introduction

Diabetes mellitus (Diabetes Mellitus, DM) is one of the common chronic diseases in the world. According to the International Diabetes Federation (International Diabetes Federation, IDF), there were about 366 million DM patients worldwide in 2021, and it is expected to increase rapidly to 783 million by 2045^[1]^. The prevalence rate of DM in China has increased rapidly in recent decades, from 0.67% in the 1980s to 12.8% in 2017, with a total of 130 million, with the highest prevalence rate of DM among countries in the world, and has become an important public health problem in China^[2**Error! Reference source not found**.^. Diabetic peripheral neuropathy (DPN) is one of the most common chronic complications of type 2 diabetes, with an incidence rate of more than 90%. Clinical manifestations range from mild paresthesia to severe ulcers, infection, muscle atrophy, neuralgia, neuropathy, neurodegenerative fracture, Charcot joint disease, etc., which are the main risk factors for foot ulcer, infection, gangrene and even amputation in diabetic patients. As the course of DPN progresses, peoples ability to work will eventually be seriously affected, reducing the quality of daily life and bringing serious financial burden to individuals and families.

Vitamin D is a steroid hormone in two main forms, vitamin D2 (ergoocalciferol) and vitamin D3 (cholecalciferol). Previous studies have found that nuclear receptors for vitamin D exist in neurons and glial cells, and that vitamin D is involved in the synthesis of neurotrophic factors and neurotransmitter synthetic enzymes^[2]^. Thus, vitamin D can delay the progression of DPN in many ways. When vitamin D levels are reduced, the risk of DPN increases in diabetes patients. A large-scale clinical study abroad found that about 81% of patients with diabetes have vitamin D deficiency, and vitamin D deficiency is closely related to the occurrence of DPN in these patients^[3]^∘

This study further explored the correlation between 25 (OH) D and DPN by observing the serum level of 25 (OH) D in patients with type 2 diabetes, aiming to provide clinical ideas for the prevention and treatment of DPN.

## 2. Research objects and methods

### 2.1 Study subjects

From January 2022 to November 2022, I visited the Second Department of Endocrinology and Metabolism, Zhangzhou Hospital, Fujian Province, Inpatient ^[5]^ who met the 1999 WHO diagnostic criteria for diabetes, T2DM, Exclusion criteria: (1) acute complications of diabetes mellitus or severe diabetic foot, acute or chronic inflammatory diseases; (2) Other types of diabetes mellitus: type 1 diabetes mellitus; Special type of diabetes mellitus; gestational diabetes mellitus; (3) Combined with other factors affecting neuropathy, such as severe spinal disease, severe liver and kidney disease, serious tumors, genetic / metabolic diseases, lack of nutrition, connective tissue diseases, trauma, neuromuscular diseases, etc.; (4) Recent administration of drugs that can cause peripheral nerve damage, Such as furazolidone, isoniazid, etc.; Long history of alcohol consumption, or exposure to toxic substances (pesticides, heavy metals); (5) Serious primary diseases such as respiratory system, circulatory system and hematopoietic system exist; (6) Mental illness; (7) incomplete data collection. A total of 210 study subjects were finally included. This study was reviewed and approved by the ethics committee of our hospital.

### 2.1 Method

General data collection: Clinical information on the study subjects, including gender, age, height, weight, waist circumference, duration of diabetes, diabetes complications, hypertension, past history, smoking history, drinking history, and body mass index (BMI) was calculated.

Detection of clinical indicators: all study subjects were fasted for 8 hours, Peripheral venous blood samples were collected by professional medical staff in the early morning of the next day, Serum 25 (OH) D, HbA1c, fasting C peptide, total cholesterol (TC), triglycerides (TG), High Density Lipoprotein Cholesterol(HDL-C), low-density lipoprotein cholesterol (LDL-C), and the blood urea nitrogen (UN), Serum Uric Acid(SUA), blood phosphorus, blood calcium, full parathyroid hormone (PTH), free triiodothyronine 3( FT3), free thyroxine 4(FT4), and the thyroid-stimulating hormone (TSH) and other indicators.

etermination of serum 25 (OH) D: using liquid chromatography-tandem mass spectrometry (liquid chromatography-tandem mass spectrometry, LC-MS / MS) determination.

Study groups: (1)serum 25 (OH) D was stratified according to the consensus on the clinical application of vitamin D and its analogues, Divided into three groups: group A (deficiency group, serum 25 (OH) D <20 ng/ml), A total of 51 cases; group B (insufficient group, 30 ng/ml> 25 (OH) D 20 ng/ml), A total of 112 cases; group C (normal group, serum 25 (OH) D 30 ng/ml), a total of 47 cases.(2)The diagnostic criteria of DPN of the Guidelines for the Prevention and Treatment of Type 2 Diabetes (2017 edition in China) were adopted: ➀ Have a clear history of diabetes; ➁Neuropathy occurring at or after the diagnosis of diabetes; ➂ The clinical symptoms of neuropathy, Such as pain, numbness, paresthesia, 5 tests (ankle reflex, vibration, pressure, temperature, acupuncture pain); ➃ If there are no clinical symptoms, Then 5 check any 2 abnormalities can also be diagnosed; ➄ Neuropathy except those resulting from other causes, If the diagnosis still cannot be confirmed according to the above examination, The need for a differential diagnosis, Neuroelectrophysiological examination can be performed. According to the above DPN diagnostic criteria, 210 selected subjects were divided into 2 groups: NDPN group: 100 cases; DPN group: 110 patients.

### 2.3 Statistical methods

Statistical analysis was performed by using the software SPSS 27.0. Distribution normality of measurement data was tested using Shapiro-Wilk. The measurement data are described by the mean standard deviation (x s) of the normal distribution, the t-test and the one-way analysis of variance, the median (interqule spacing) [M (P25, P75)] by the nonparametric Mann-Whitney U test, and the non-parametric Kruskal-Wallis H test for the mean-value between multiple groups. The number of cases (%), χ 2 test between two groups and chi-square test for pairwise comparisons between multiple groups. The correlation analysis of clinical data with 25 (OH) D level and DPN was studied by Spearman rank correlation analysis. Ordered Logistic regression was used to analyze the relationship between 25 (OH) D levels and multiple influencing factors. Univariate and multivariate binary Logistic regression analysis was used to explore the main risk factors for DPN. The receiver operating characteristic curve (ROC) was calculated for the maximum Youden index of the main risk factors to determine the optimal cutoff value. The clinical predictive value criterion is the area under the ROC curve. All tests were two-sided, P <0.05 indicates differencesSignificsignificance, 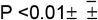

## 3. Results

### 3.1 Intergroup analysis of different serum 25 hydroxyvitamin D groups

#### 3.1.1 Comparison of general clinical data in different serum 25-hydroxyvita-min D level groups

The comparison of general data between groups A, B and C showed significant differences in blood calcium and DPN (P <0.05), gender, age, RHR, waist circumference, BMI, HBP and Hnocturnal hypoglycemia, HbA 1 c, blood phosphorus, TG, TC, HDL-C, LDL-C, BUN, TSH, FT3, FT4, FT 4, fasting C peptide, UA, PTH, macrovascular complications, DKD, and DR (P> 0.05), detailed in Table 1.

**Table 1:** The comparison of common data among different subgroups of VD.

| | Group A<br>(n= 51) | Group B<br>(n=112) | Group C<br>(n= 47) | F/ z/$\chi^2$ | P-value |
| --- | --- | --- | --- | --- | --- |
| Gender (male / female) | (30/21) | (68/44) | (24/23) | 1.281<sup>a</sup> | .527 |
| Age (year) | 52.24±13.88 | 57.02±11.11 | 58.26±11.80 | .274<sup>c</sup> | .761 |
| RHR (sub / min) | 87 (78,97) | 84 (78, 95) | 81.5 (74, 88) | 3.16<sup>d</sup> | 0.206 |
| waistline (cm) | 86 (21.51,25.26) | 85 (21.51, 25.26) | 85 (21.51 25.26) | 0.717<sup>d</sup> | 0.410 |
| BMI (kg/m<sup>2</sup>) | 23.43 (22.30,25.20) | 23.47 (21.48,24.78) | 23.62 (22.22,25.39) | 0.420<sup>d</sup> | 0.811 |
| HBP [Case number (%)] | 27 (52.9%) | 45(40.2%) | 26 (55.3%) | 3.665<sup>a</sup> | 0.160 |
| Low blood sugar at night<br>[Number of cases, (%)] | 9 (17.6%) | 12 (10.7%) | 2 (4.3%) | 4.34 1<sup>a</sup> | 0.114 |
| HbA1c (%) | 10.35 (7.72,12.23) | 9.42 (7.72,11.54) | 12.29 (8.43, 12.48) | 0.037<sup>d</sup> | 0.982 |
| Blood phosphorus (mmol/L) | 1.30 (1.15, 1.48) | 1.27 (1.12, 1.49) | 1.26 (1.09, 1.49) | 2.073<sup>d</sup> | 0.355 |
| Blood calcium (mmol/L) | 2.22 (2.17, 2.29) | 2.31 (2.23, 2.38) | 2.32 (2.22, 2.39) | 10.710<sup>d</sup> | 0.005** |
| TG (mmol/L) | 1.26 (1.01, 1.72) | 1.43 (1.04, 2.48) | 1.85 (1.42, 2.48) | 4.404<sup>d</sup> | 0.111 |
| TC (mmol/L) | 4.72 (3.72, 5.56) | 4.78 (4.10, 5.72) | 4.86 (3.86, 5.59) | 1.396<sup>d</sup> | 0.498 |
| HDL-C (mmol/L) | 1.03 (0.89, 1.24) | 1.09 (0.91, 1.28) | 0.93 (0.86, 1.10) | 2.043<sup>d</sup> | 0.360 |
| BUN (mmol/L) | 5.50 (4.30, 7.40) | 5.4 (4.70, 6.60) | 5.50 (4.50, 7.30) | 0.216<sup>d</sup> | 0.898 |
| TSH (pmol/L) | 1.55 (1.13, 2.25) | 1.38 (0.79, 2.11) | 1.71 (0.94, 2.73) | 2.529<sup>d</sup> | 0.282 |
| FT3 (pmol/L) | 4.72 (4.30, 5.18) | 4.85 (4.40, 5.30) | 4.94 (4.41, 5.40) | 1.522<sup>d</sup> | 0.467 |
| FT4 (pmol/L) | 11.60 (10.10, 13.3) | 11.70 (10.80, 13.45) | 11.65 (10.20, 14.80) | 0.703<sup>d</sup> | 0.703 |
| Fasting C-peptide (ng/mL) | 0.76 (0.50, 1.06) | 0.77 (0.56, 1.32) | 1.11 (0.63, 1.69) | 2.600<sup>d</sup> | 0.273 |
| UA (umol/L) | 311.22±112.22 | 323.97 ±90.63 | 323.21 ±126.59 | .275<sup>c</sup> | 0.76 |
| PTH(pg/mL) | 41.30 (32.30, 49.90) | 33.90 (27.05, 41.60) | 37.05 (23.30,52.70) | 2.462<sup>d</sup> | 0.297 |
| DPN (Case %) | 36 (70.6%) | 54 (47.8%) | 20 (42.6%) | 9.378<sup>a</sup> | .009** |
| Complications of large vessels<br>(Example%) | 30 (58.8%) | 72 (64.3%) | 28 (59.6%) | .089<sup>a</sup> | .956 |
| DKD (Case %) | 11 (21.6%) | 22 (19.6%) | 12 (25.5%) | .857<sup>a</sup> | .651 |
| DR (Case %) | 13 (25.5%) | 21 (18.6%) | 10 (21.3%) | 1.149<sup>a</sup> | .563 |

Data are expressed as mean standard deviation, median (interquartile spacing), and number of cases (%).<sup>a</sup> is $\chi^2$ Values, <sup>c</sup> is the F value, <sup>d</sup> is the z-value.\* P <0.05, with a statistically significant difference,\*\* P <0.01, indicating a statistically significant difference

#### 3.1.2 Correlation analysis of serum 25 hydroxyvitamin D and various factors

According to the comparison of different serum 25 hydroxyvitamin D levels with clinical general data, laboratory data and characteristic data, the correlation between serum 25 hydroxyvitamin D and the above statistically significant factors was analyzed. The results showed that serum 25 hydroxyvitamin D was positively associated with blood calcium (r=0.224, P <0.0.01), and serum 25 hydroxyvitamin D was negatively associated with DPN (r= -0.0195, P <0.005) (P <0.05). The results suggested a strong correlation between serum 25 hydroxyvitamin D and DPN. See Table 2 for details.

**Table 2.**
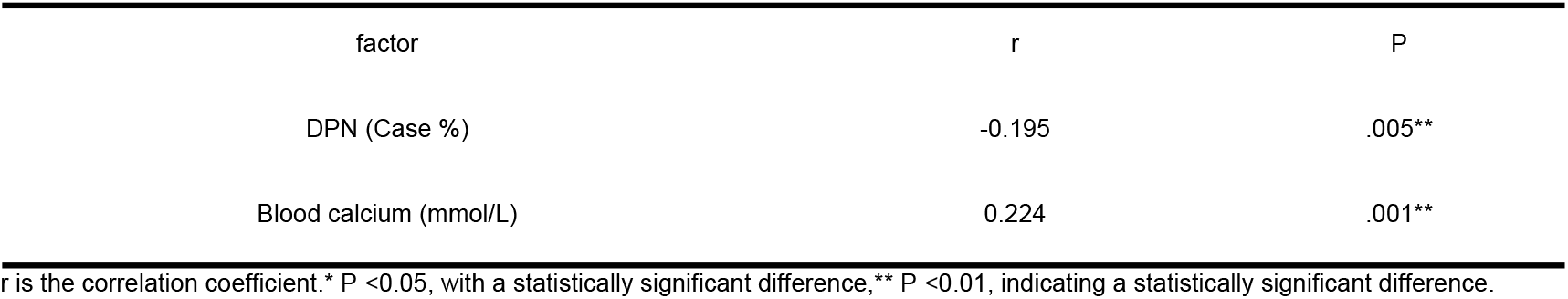
Analysis of the association of VD with various factors.

#### 3.1.3 Orderly Logistic regression analysis of serum 25 hydroxyvitamin D and related influencing factors

After different serum 25 hydroxyvitamin D levels as the dependent variable, the significant associated with different serum 25 hydroxyvitamin D levels: serum calcium and DPN as independent variables, the last retained factors were blood calcium and DPN (P <0.05), and the results showed that DPN (OR = 2.100,95% CI 0.224∼1.301, P=0.006), blood calcium (OR = 1.048,95% CI 0.486 to 3.755, P=0.011) were independently associated with serum 25 hydroxyvitamin D, as detailed in Table 3.

**Table 3.** Ordered logistic regression analysis of different VD subgroups.

| correlative factor | $\beta$ | S.E. | Wald $\chi^2$ | P | OR (95% CI) |
| --- | --- | --- | --- | --- | --- |
| DPN | 0.763 | 0.275 | 7.691 | .006** | 2.100 ( 0.224, 1.301) |
| Blood calcium (mmol/L) | 2.120 | 0.834 | 6.491 | .011* | 1.048 ( 0.486, 3.755) |

OR: odds ratio; 95% CI: 95% confidence interval.\* P <0.05, a statistically significant difference,\*\* P <0.01, indicating a statistically significant difference.

### 3.2 Intergroup analysis between the NDPN and DPN groups

#### 3.2.1 Comparison of general data between NDPN and DPN groups

Comparison of general data between NDPN and DPN groups showed statistically significant differences in BMI, RHR, TG, FT3, BUN, 25-hydroxyvitamin D3, total 25-hydroxyvitamin D (P <0.05); gender, age, HBP, history of nighttime hypoglycemia, HbA 1 c, blood phosphorus, calcium, TC, HDL-C, LDL-C, TSH, FT4, PTH, fasting C peptide, UA (P> 0.05). Both the BMI and RHR levels were higher in the DPN group than those in the NDPN group. See Table 4 for details.

**Table 4.**
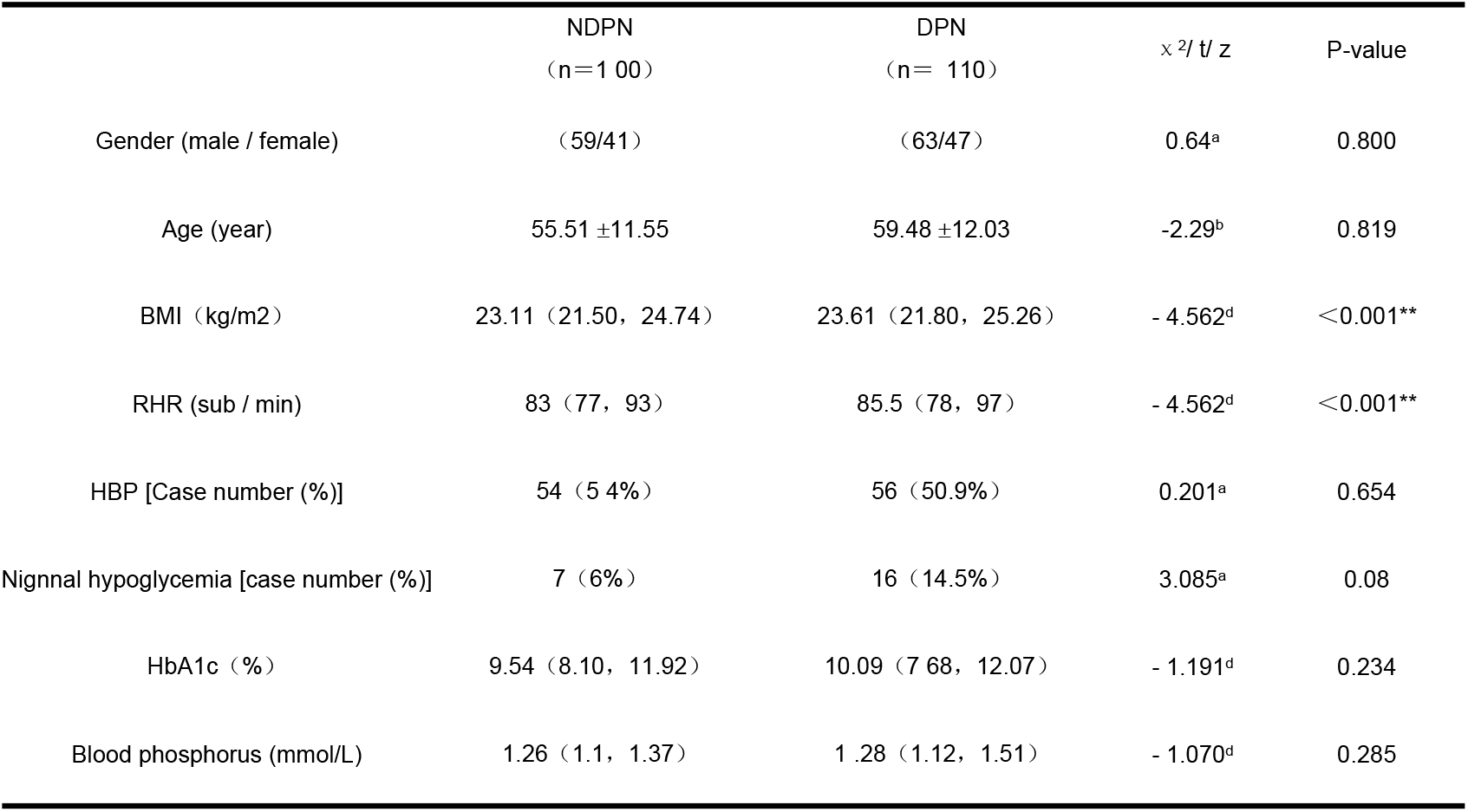
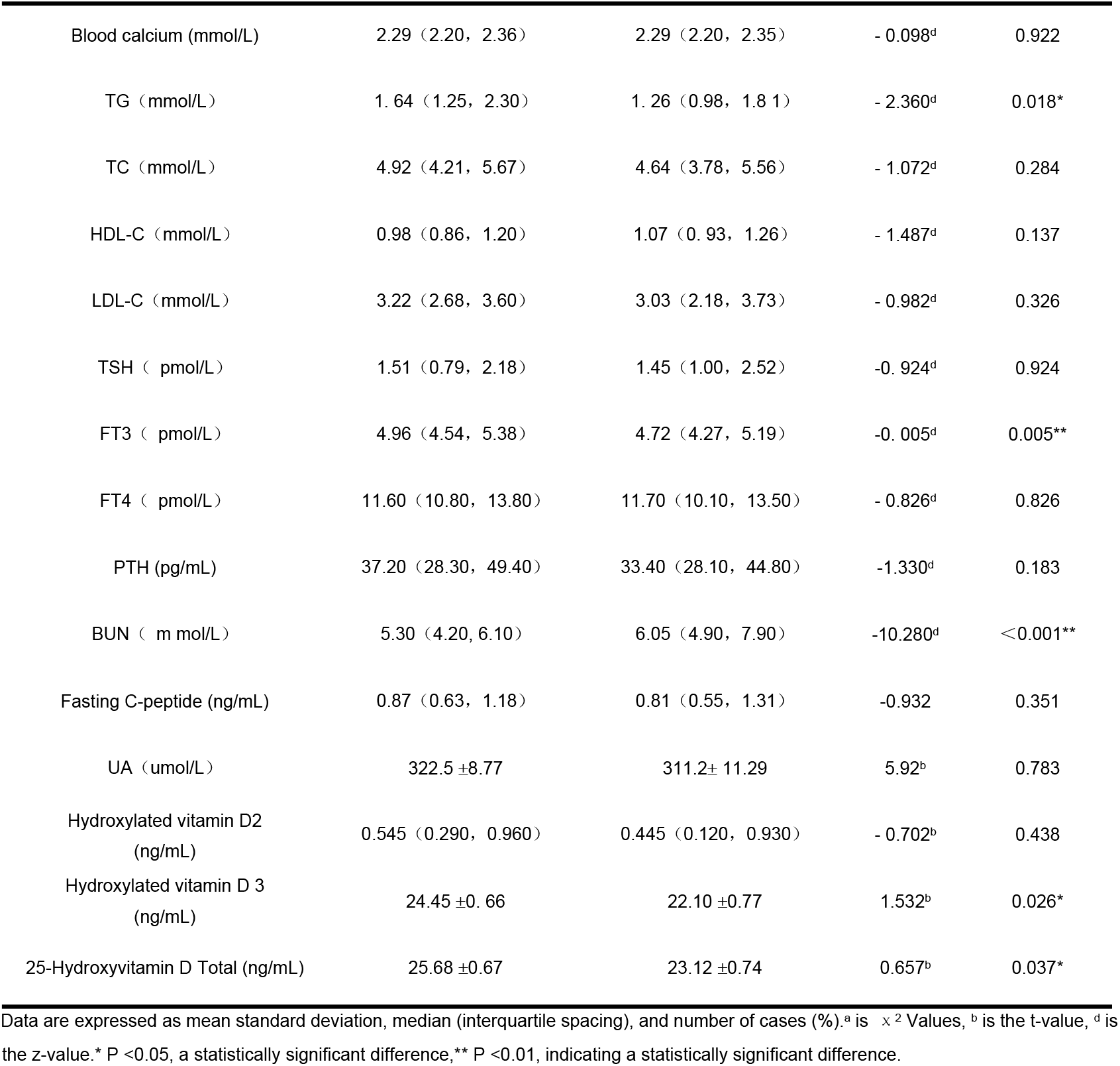
The comparison of common data between NPN and DPN.

| | NDPN<br>(n=100) | DPN<br>(n=110) | $\chi^2/t/z$ | P-value |
| --- | --- | --- | --- | --- |
| Gender (male / female) | (59/41) | (63/47) | 0.64<sup>a</sup> | 0.800 |
| Age (year) | 55.51 ±11.55 | 59.48 ±12.03 | -2.29<sup>b</sup> | 0.819 |
| BMI (kg/m<sup>2</sup>) | 23.11 (21.50, 24.74) | 23.61 (21.80, 25.26) | -4.562<sup>d</sup> | <0.001** |
| RHR (sub / min) | 83 (77, 93) | 85.5 (78, 97) | -4.562<sup>d</sup> | <0.001** |
| HBP [Case number (%)] | 54 (54%) | 56 (50.9%) | 0.201<sup>a</sup> | 0.654 |
| Nocturnal hypoglycemia [case number (%)] | 7 (6%) | 16 (14.5%) | 3.085<sup>a</sup> | 0.08 |
| HbA1c (%) | 9.54 (8.10, 11.92) | 10.09 (7.68, 12.07) | -1.191<sup>d</sup> | 0.234 |
| Blood phosphorus (mmol/L) | 1.26 (1.1, 1.37) | 1.28 (1.12, 1.51) | -1.070<sup>d</sup> | 0.285 |
| Blood calcium (mmol/L) | 2.29 (2.20, 2.36) | 2.29 (2.20, 2.35) | - 0.098<sup>d</sup> | 0.922 |
| TG (mmol/L) | 1.64 (1.25, 2.30) | 1.26 (0.98, 1.81) | - 2.360<sup>d</sup> | 0.018* |
| TC (mmol/L) | 4.92 (4.21, 5.67) | 4.64 (3.78, 5.56) | - 1.072<sup>d</sup> | 0.284 |
| HDL-C (mmol/L) | 0.98 (0.86, 1.20) | 1.07 (0.93, 1.26) | - 1.487<sup>d</sup> | 0.137 |
| LDL-C (mmol/L) | 3.22 (2.68, 3.60) | 3.03 (2.18, 3.73) | - 0.982<sup>d</sup> | 0.326 |
| TSH (pmol/L) | 1.51 (0.79, 2.18) | 1.45 (1.00, 2.52) | -0.924<sup>d</sup> | 0.924 |
| FT3 (pmol/L) | 4.96 (4.54, 5.38) | 4.72 (4.27, 5.19) | -0.005<sup>d</sup> | 0.005** |
| FT4 (pmol/L) | 11.60 (10.80, 13.80) | 11.70 (10.10, 13.50) | - 0.826<sup>d</sup> | 0.826 |
| PTH (pg/mL) | 37.20 (28.30, 49.40) | 33.40 (28.10, 44.80) | -1.330<sup>d</sup> | 0.183 |
| BUN (mmol/L) | 5.30 (4.20, 6.10) | 6.05 (4.90, 7.90) | -10.280<sup>d</sup> | <0.001** |
| Fasting C-peptide (ng/mL) | 0.87 (0.63, 1.18) | 0.81 (0.55, 1.31) | -0.932 | 0.351 |
| UA (μmol/L) | 322.5 ± 8.77 | 311.2 ± 11.29 | 5.92<sup>b</sup> | 0.783 |
| Hydroxylated vitamin D2 (ng/mL) | 0.545 (0.290, 0.960) | 0.445 (0.120, 0.930) | - 0.702<sup>b</sup> | 0.438 |
| Hydroxylated vitamin D3 (ng/mL) | 24.45 ± 0.66 | 22.10 ± 0.77 | 1.532<sup>b</sup> | 0.026* |
| 25-Hydroxyvitamin D Total (ng/mL) | 25.68 ± 0.67 | 23.12 ± 0.74 | 0.657<sup>b</sup> | 0.037* |

Data are expressed as mean standard deviation, median (interquartile spacing), and number of cases (%).<sup>a</sup> is $\chi^2$ Values, <sup>b</sup> is the t-value, <sup>d</sup> is the z-value. \* P < 0.05, a statistically significant difference, \*\* P < 0.01, indicating a statistically significant difference.

#### 3.2.2 Correlation analysis between DPN and each factor

According to the results of the comparison between the general data, laboratory data, serum 25 hydroxyvitamin D level and other factors between the NDPN and DPN groups, DPN and statistically significant factors between the above groups, The results indicate that: DPN and BUN (r=0.869, P<0.001), RHR(r=0.579, P <0.001) showed a positive correlation; Compared with 25-hydroxyvitamin D3 (r= -0.192, P=0.005), 25-hydroxyvitamin D total (r= -0.192, P=0.005), FT3(r=-0.202, P=0.004), TG(r=-0.164, P=0.018), BMI(r=-0.709, P <0.001) showed a negative correlation. Among them, DPN showed a strong correlation with 25-hydroxyvitamin D3,25-hydroxyvitamin D total, FT3, BMI, BUN, and RHR, as detailed in Table 5.

**Table 5.** Association analysis of D PN with the related factors.

|  | r | P |
| --- | --- | --- |
| Hydroxylated vitamin D3 (ng/mL) | -0.192 | 0.005** |
| Total of hydroxylated vitamin D (ng/mL) | -0.192 | 0.005** |
| FT3 (pmol/L) | -0.202 | 0.004** |
| TG (mmol/L) | - 0.164 | 0.018* |
| BUN (mmol/L) | 0.869 | p<0.001** |
| BMI (kg/m<sup>2</sup>) | -0.709 | p<0.001** |
| RHR (sub / min) | 0.579 | p<0.001** |

r is the correlation coefficient.\* P <0.05, a statistically significant difference,\*\* P <0.01, indicating a statistically significant difference.

#### 3.2.3 Logistic regression analysis of DPN and influencing factors

Univariate binary Logistic regression analysis was conducted one by one with the occurrence of DPN as the dependent variable and other clinical indicators as the independent variables. The results showed that 25-hydroxyvitamin D3 (OR = 0.944, 95% CI 0.908 to 0.981, P=0.003), 25-hydroxyvitamin D total (OR = 0.944,95% CI 0.907∼0.981,P=0.004)、FT3(OR=0.566, 95% CI 0.379∼ 0.845,P=0.005)、BMI(OR=0.119, 95% CI 0.70∼0.200,P<0.001)、RHR(OR=1.160,95%m CI 1.115∼1.206, P <0.001) may be a risk factor for predicting the occurrence of DPN (P <0.05), as detailed in Table 6.

**Table 6.** Univariate binary logistic regression analysis of DPN.

| correlative factor | $\beta$ | S.E. | Wald | P | OR (95% CI) |
| --- | --- | --- | --- | --- | --- |
| Hydroxylated vitamin D 3 (ng/mL) | -0.58 | 0.20 | 8.601 | 0.003** | 0.944 (0.908, 0.981) |
| Total of hydroxylated vitamin D (ng/mL) | -0.58 | 0.20 | 8.377 | 0.004** | 0.944 (0.907, 0.981) |
| FT3 (pmol/L) | -0.569 | 0.205 | 7.729 | 0.005** | 0.566 (0.379, 0.845) |
| TG (mmol/L) | -0.257 | 0.132 | 3.796 | 0.051 | 0.774 (0.598, 1.002) |
| BMI (kg/m<sup>2</sup>) | -2.132 | 0.267 | 63.596 | <0.001** | 0.119 (0.70, 0.200) |
| RHR (sub / min) | 0.148 | 0.20 | 55.115 | <0.001** | 1.160 (1.115, 1.206) |
| BUN (mmol/L) | 46.920 | 1472.705 | 0.001 | 0.975 |  |

OR: odds ratio; 95% CI: 95% confidence interval.\* P <0.05, with a statistically significant difference,\*\* P <0.01, indicating a statistically significant difference.

Taking the occurrence of DPN as the dependent variable and including the above risk factors as the independent variables, a multivariate binary Logistic regression model was established. After adjusting for confounding factors, the final results showed that the total 25-hydroxyvitamin D (OR = 0.935,95% CI 0.893∼0.978,P=0.004)、FT3(OR=0.590,95% CI 0.387∼0.900, P=0.014) was a protective factor of DPN (P <0.05)(Table 7)

**Table 7.** Multivariate binary logistic regression analysis of DPN.

| correlative factor | $\beta$ | S.E. | Wald | P | OR (95% CI) |
| --- | --- | --- | --- | --- | --- |
| Total of hydroxylated vitamin D (ng/mL) | -0.067 | 0.023 | 8.492 | 0.004** | 0.935 (0.893, 0.978) |
| FT3 (pmol/L) | -0.527 | 0.216 | 5.985 | 0.014* | 0.590 (0.387, 0.900) |
| BMI (kg/m<sup>2</sup>) | 0.008 | 0.019 | 0.186 | 0.666 | 1.008 (0.971, 1.046) |
| RHR (sub / min) | 0.019 | 0.011 | 2.806 | 0.094 | 1.019 (0.997, 1.041) |

OR: odds ratio; 95% CI: 95% confidence interval.\* P <0.05, with a statistically significant difference,\*\* P <0.01, indicating a statistically significant difference.

Taking the occurrence of DPN as the dependent variable and including the above risk factors as the independent variable, a multivariate binary Logistic regression model was established. After adjusting for confounding factors, the final results showed that 25-hydroxyvitamin D3 (OR = 0.944, 95% CI 0.906∼0.984,P=0.007)、FT3(OR=0.593, 95% CI 0.388 to 0.905, P=0.015) was a protective factor for DPN (P <0.05)(Table 8).

**Table 8.** Multivariate binary logistic regression analysis of DPN.

| correlative factor | $\beta$ | S.E. | Wald | P | OR (95% CI) |
| --- | --- | --- | --- | --- | --- |
| Hydroxylated vitamin D 3 (ng/mL) | -0.057 | 0.021 | 7.302 | 0.007** | 0.944 (0.906, 0.984) |
| FT3 (pmol/L) | -0.523 | 0.216 | 5.865 | 0.015* | 0.593 (0.388, 0.905) |
| BMI (kg/m<sup>2</sup>) | 0.008 | 0.019 | 0.198 | 0.656 | 1.008 (0.972, 1.046) |
| RHR (sub / min) | 0.019 | 0.11 | 2.975 | 0.085 | 1.020 (0.997, 1.042) |

OR: odds ratio; 95% CI: 95% confidence interval.\* P <0.05, with a statistically significant difference,\*\* P <0.01, indicating a statistically significant difference.

#### 3.2.4 ROC curve analysis

The results of receiver operating characteristic curve (ROC curve) analysis showed that the optimal cutoff value of 25-hydroxyvitamin D3 predicting DPN in T2DM was 18.85ng/ml, with AUC of 0.76, sensitivity of 91% and specificity of 48.2%; the optimal cutoff value of 25-hydroxyvitamin D patients predicting T2DM was 19.94ng/ml, AUC was 0.765, sensitivity was 90%, and specificity was 50% respectively(Table 9,Figure1).

**Table 9.** ROC analysis of VD3(25-OH)、VD(25-OH) predicting DPN.

|  | AUC | P | 95% CI |  | Best threshold | sensitivity | specificity | Yorden index |
| --- | --- | --- | --- | --- | --- | --- | --- | --- |
| 25-hydroxy vitamin D3 | 0.76 | p< 0.001** | 0.697 | 0.823 | 18.85 | 91% | 48.2% | 0.392 |
| 25-hydroxy vitamin D | 0.765 | p< 0.001** | 0.703 | 0.828 | 19.94 | 90.0% | 50.0% | 0.4 |

95% CI: 95% confidence interval.\* P <0.05, with a statistically significant difference,\*\* P <0.01, indicating a statistically significant difference.

**Figure 1.**
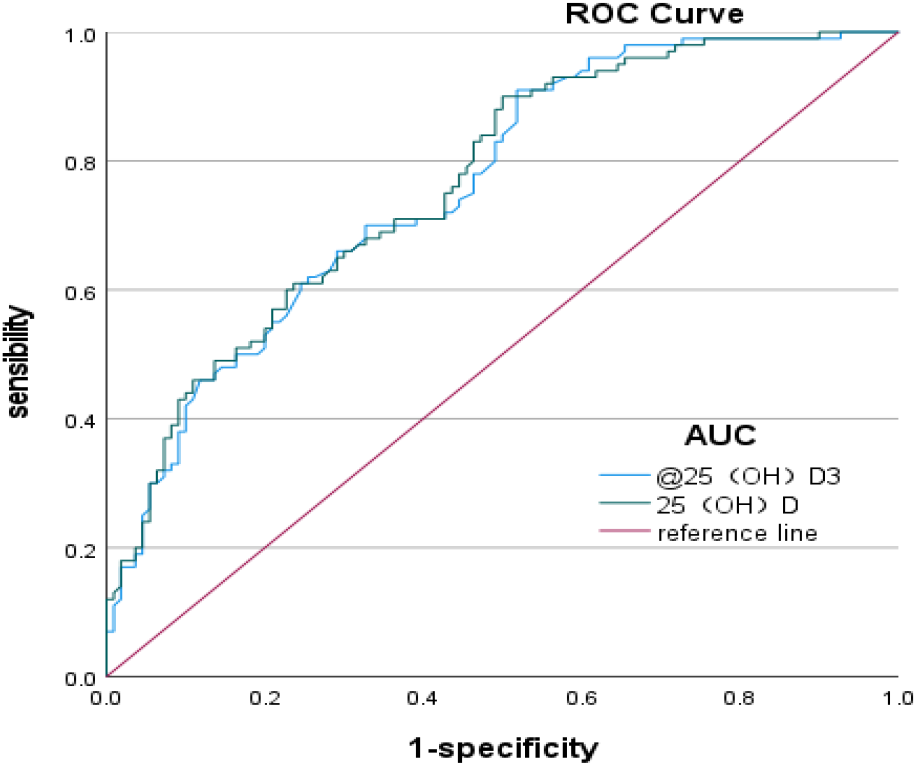
ROC curves of total 25 H D and 25 H D3 predicting the occurrence of DCAN

## 4. Discuss

Type 2 diabetes mellitus (T2DM), a complex metabolic disease affecting both developing and developed countries, is reported to be increasing at an alarming rate^[6]^.With the rapid development of economy and the acceleration of industrialization, the change of lifestyle and the acceleration of the aging process, the prevalence of diabetes in China has risen rapidly. It has become an important chronic non-communicable disease that seriously endangers peoples health after cardiovascular diseases and tumors. According to the World Health Organization, Chinas economic losses from 2005 to 2015 due to diabetes and related cardiovascular diseases reached us $557.7 billion^[7]^.The global prevalence of diabetes has risen dramatically, reaching 8.3% in 2014, which corresponds to 3.87 million patients^**Error! Reference source not found**.^.In the early stage of the disease, it can only show dry mouth, polydipsia, polyuria or weight loss, which does not affect daily life, and often does not cause attention; with the extension of the disease, poor blood glucose control and long-term metabolic disorders, often cause chronic microvascular complications such as nerve, eye and kidney lesions, as well as induce cardiovascular and cerebrovascular diseases. Diabetic microvascular complications include diabetic neuropathy, diabetic retinopathy, diabetic nephropathy, and mostly occur in patients with a course of diabetes for several years or decades^[8]^; Type 2 diabetes is associated with systemic chronic inflammation characterized by increased concentrations of acute phase response proteins and inflammatory markers, these inflammatory mediators that may contribute to insulin resistance and β -cell dysfunction^[9][10]^, Because vitamin D has anti-inflammatory and immunomodulatory effects, it can ameliorate diabetic low-grade chronic inflammation by regulating cytokine production^[11]^; The main pathogenesis of type 2 diabetes for insulin resistance, vitamin D can not only by regulating the insulin receptor gene expression to increase insulin secretion, also can by restoring the damaged islet cells and stimulate islet β cells increase insulin secretion, reduce β cell dysfunction, so as to improve the insulin sensitivity of type 2 diabetes patients, better control blood sugar^[12]^. Therefore, vitamin D has a protective effect on type 2 diabetes, and vitamin D deficiency can lead to an increased risk of type 2 diabetes^[13]^∘

Diabetic peripheral neuropathy (DPN) is one of the most common chronic complications of diabetes mellitus. DPN affects a wide range and combines different degrees of DPN^[14]^.There is usually a risk of increased physical disability, cardiovascular disease, and mortality. At least 20% of patients with type 1 diabetes develop distal symmetric polyneuropathy within 20 years, and 10% to 20% of newly diagnosed type 2 diabetic patients with distal symmetric polyneuropathy increase to 50% within 10 years.**Error! Reference source not found**.The associated neurological complications place a huge burden on the patient and on society as a whole^[15]^.DPN is mainly due to decreased quality of life with pain, sensory loss, gait instability, foot ulceration, and amputation^[16]^.Furthermore, the occurrence and development of these complications can lead to loss of vision and neurological function, impaired activity and cognition, reduced quality of life, limited employment and productivity, and increased costs for patients and society. Without control or treatment, irreversible damage or even death may occur^[17]^.In the occurrence and development of DPN put forward a variety of possible causes, in the past decades, mainly believe that long-term hyperglycemia state cause polyol bypass pathway, oxygen radical generation, late glycation terminal products, inflammation, microcirculation perfusion, oxidative stress response, direct damage or indirect harmful to nerve. In recent years, studies have found that in addition to glycemic factors, age-related neuronal wear, hypertension, blood lipid levels, body weight, and decreased neurotrophic factors interact to lead to the occurrence and development of peripheral neuropathy. Later, some studies found that vitamin D level is significantly reduced in DPN. It is speculated that vitamin D is a risk factor for DPN and may be involved in the development of DPN, such as oxidative stress caused by hyperglycemia, inflammation and neuronal ischemia. The mechanism of DPN is still incompletely understood, and besides strict glycemic control, an effective modified therapy of DPN disease is lacking^[18][19]^. The identification of DPN risk factors is crucial for a better understanding of the mechanisms of DPN and more effective treatments.

Vitamin D is similar to vitamins A, E, and K, and it is A fat-soluble vitamin that exists in different forms: vitamin D3 (cholecalciferol) and vitamin D2. Vitamin D3 is synthesized in the skin under UV B (UV-B) irradiation, while vitamin D2 comes from the diet and is converted to ergocalciferol by UV B. In the liver, both cholecalciferol and ergogalciferol are converted to 25-hydroxylated vitamins D2 and D3. It is then converted in the kidney to the active form by 1-α-hydroxylase, namely 1,25-dihydroxyvitamin D^[20]^. 44 It is the main form of VD in the blood circulation, the most abundant and most stable form of serum VD metabolites, and is the main storage form of VD in the body. For its good stability and long plasma half-life, 25 (OH) D was used as a gold index to evaluate serum vitamin D levels in medical studies^[21]^. In this study, serum [25- (OH) D] level was determined by most common clinical detection methods in China, mass spectrometry, including serum [25- (OH) D], serum [25- (OH) D3], and serum [25- (OH) D2], which were compared as the study index and 25 (OH) D as the main study index.

In addition to participating in bone metabolism, serum vitamin D also participates in the development of a variety of diseases in the body, such as cardiovascular diseases, metabolic diseases, tumors, multiple sclerosis, biological infections and autoimmune diseases^[22]^, And also involved in the proliferation and differentiation of immune cells and other immune regulation of the body^[23]^. Vitamin D deficiency is prevalent worldwide and affects from 30% to 87% of the population. D Vitamin D deficiency is usually defined as a 25 (OH) D concentration of 2030 ng / ml (5075 nmol / l), and vitamin D deficiency is defined as a 25 (OH) D concentration of less than 20 ng / ml (<50 nmol / l). Vitamin D regulates insulin secretion by binding to the vitamin D receptors present on pancreatic β (B) cells^[24]^. Vitamin D increases insulin sensitivity by increasing insulin receptor expression by binding to the vitamin D response element present in the promoter of the human insulin receptor gene^[25][26]^. It also affects fatty acid metabolism in insulin-responsive tissues through activation of its transcription factors^[27]^, And is protective against cytokine-induced apoptosis^[28][29][30]^. Thus, there is an inverse relationship between vitamin D and diabetic complications.

To investigate the association between DPN and 25-hydroxyvitamin D, According to the diagnostic criteria of DPN, this study divided patients with type 2 diabetes into DPN and NDPN groups, Comparison of the general data and the biochemical indicators between the two groups, Showed that serum 25 (OH) D and 25 (OH) D3 were decreased in both DPN and NDPN, And the mean level of 25 (OH) D of (23.12±0.74ng/ml) in the DPN group was significantly lower than that in the NDPN group (25.68±0.67ng/ml), The mean level of 25 (OH) D3 was (22.10±0.77ng/ml) was significantly lower than that in the NDPN group (24.45±0.66ng/ml), DPN, 81.8% decreased the serum 25 (OH) D, 25 (OH) D3 of the patients in the group, In contrast to 73% in the NDPN group, Thus, 25 (OH) D reduction is more common in DPN. In an observational study of T2DM, patients over 5 years showed a significant reduction in their 25 (OH) D levels. This suggests that low serum vitamin D level is not only a relevant risk factor for DPN, but also a new therapeutic to treat DPN. In addition, studies have shown that vitamin D is negatively correlated with the presence and severity of DPN. With the increase of serum vitamin D level, the severity of nerve conduction velocity decreases. After 3 months after vitamin D supplementation treatment, the symptoms of neuropathy pain are reduced by about 50%^[31]^. Serum 25 (OH) D was stratified according to the Consensus on the clinical Application of Vitamin D and its analogues, respectively: group A (deficiency group, serum 25 (OH) D <20 ng/ml); group B (insufficient group, 30 ng/ml> serum 25 (OH) D 20 ng/ml); group C (normal group, serum 25 (OH) D>30ng / ml); the incidence of peripheral neuropathy in the three groups was 70.6%, 47.8% and 42.6%, respectively. With the increase of serum 25 (OH) D level, the incidence of diabetic peripheral neuropathy gradually decreased; using the Logistic logistic regression analysis, observing the effect of 25 (OH) D3, 25 (OH) D, FT3, TG, BMI, RHR, BUN on DPN, the results showed that serum 25 (OH) D and FT3 were the protective factors of DPN in type 2 diabetes.

The receiver operating characteristic curve showed that the optimal cutoff for 25 (OH) D3 for DPN in T2DM was 18.85ng/ml (91% sensitivity, with 48.2% specificity) with an AUC of 0.76; the optimal cutoff for 25 (OH) D for T2DM was 19.94ng/ml (sensitivity 90%, 50% specificity) with an AUC of 0.765.

The results showed that T2DM patients with lipid disorders (especially hypertriglyceridemia) were more prone to DPN, even without T2DM, hypertriglyceridemia was still significantly associated with peripheral neuropathy, patients with high TG increased 2.1 times DPN, an important independent risk factor for DPN, especially in small unmyelinated axons^[32]^. However, in our study, between the NDPN group and the NDPN group, the high TG level in the NDPN group, considering the patients recent poor blood glucose control, taking lipid-lowering drugs, different dietary structure, smoking, alcohol consumption and other interfering factors may affect the study results.

In addition, this study also found that FT3 is a protective factor of DPN, 3,5,3′triiodome-thyronine (3,5,3′-triiodothyronine, T3) can inhibit the axonal expansion of sensory neurons, stimulate new axons to increase the number of active neurons, and get nerve repair and regeneration. Previous animal experiments have found that T3 can promote the growth of dorsal root ganglia and simple neurons. And in the chronic hyperglycemia state, T3 decline will lead to vascular endothelial dysfunction, resulting in insufficient nerve blood supply, and thus damage to the peripheral nerves. Thus, thyroid hormones can protect microvascular endothelial cells, thereby protecting peripheral nerves from hyperglycemia.

### 4.1 Study limitations

The study has the following deficiencies: 1. Using retrospective study design, non-diabetic population was not included as a control group.2, a small sample size.3. No long-term follow-up study was conducted, and the change of 25 (OH) D before and after DPN treatment was further counted to assess whether it could be used as an indicator to help determine the efficacy; 4. No further statistics were made on patients taking calcium tablets, sunshine and diet, and the influence of diet, environment and drug differences on 25 (OH) D cannot be excluded.

### 5. Conclusion

The reduction of 25 (OH) D level in T2DM patients is closely related to the occurrence of DPN. 25 (OH) D is a protective factor of DPN, especially when the level of 25 (OH) D is <19.94ng/ml, the occurrence of DPN should be vigilant. Testing of 25 (OH) D is helpful for the early diagnosis of DPN and can provide an objective basis for the early intervention of DPN.

## Data availability statement

The original contributions presented in the study are included in the article/supplementary material. Further inquiries can be directed to the corresponding authors.

## Ethics statement

The studies involving humans were approved by the ethics committee of Zhangzhou Affiliated Hospital of Fujian Medical University.

## Publisher’s note

All claims expressed in this article are solely those of the authors and do not necessarily represent those of their affiliated organizations, or those of the publisher, the editors and the reviewers. Any product that may be evaluated in this article, or claim that may be made by its manufacturer, is not guaranteed or endorsed by the publisher.

